# Comorbidity alters the genetic relationship between anxiety disorders and major depression

**DOI:** 10.1101/2024.11.19.24317523

**Authors:** Markos Tesfaye, Alexey Shadrin, Nadine Parker, Piotr Jaholkowski, Pravesh Parekh, Gleda Kutrolli, Viktoria Birkenæs, Nora R. Bakken, Helga Ask, Guy Hindley, Oleksandr Frei, Srdjan Djurovic, Anders M. Dale, Olav B. Smeland, Kevin S. O’Connell, Ole A. Andreassen

**Author notes:** **Corresponding authors**: Markos Tesfaye, M.D., Ph.D. and, Ole Andreassen, M.D., Ph.D., Division of Mental Health and Addiction, Oslo University Hospital & Institute of Clinical Medicine, University of Oslo, Building 49, Oslo University Hospital, Ullevål, Kirkeveien 166, PO Box 4956 Nydalen, 0424 Oslo, Norway.

## Abstract

**Background:** The extensive genetic overlap between anxiety disorders (ANX) and major depression (MD) may partly reflect inclusion of comorbid cases in genome-wide association studies (GWAS). We investigated this genetic relationship between ANX and MD with and without mutual comorbidity.

**Methods:** Using UK Biobank, we performed disorder-specific GWAS for ANX-only (cases/controls = 9,980/179,442) and MD-only (cases/controls = 15,301/179,038) and derived polygenic risk scores (PRS). In the Norwegian Mother, Father, and Child Cohort (MoBa), we tested associations of PRS with MD-only (N=7,486), ANX-only (N=1,992), and comorbid (ANX-MD) (N=3,468) and controls (N=85,851). PRS associations with anxiety and depression symptoms were tested in MoBa (N=54,862). GWAS including comorbid cases (MD-comorbid or ANX-comorbid) were used for comparison. Genetic correlations were compared by comorbidity status, and Mendelian randomization was employed to assess causal relationships.

**Results:** MD-comorbid and ANX-comorbid PRS showed stronger association with ANX-MD cases than with their primary disorders, MD-only (Z=-2.82; P_adjusted_=0.01) and ANX-only (Z=-2.36; P_adjusted_=0.03), respectively. MD-only PRS was more strongly associated with MD-only than with ANX-only cases (Z=3.63; P_adjusted_=6.9e-04). The genetic correlation was lower between ANX-only and MD-only (rg=0.53, SE=0.11) than between ANX-comorbid and MD-comorbid (rg=0.91, SE=0.01). Bidirectional causal effects observed in comorbidity-inclusive analyses were attenuated to null when comorbid states were excluded. Gene sets of MD-comorbid, ANX-comorbid, and MD-only, but not of ANX-only, were enriched for the immune regulation pathway – interleukin-21 production.

**Conclusions:** The genetic distinction between ANX and MD becomes more pronounced when comorbid cases are excluded. The findings underscore the importance of disorder-specific genetic studies for advancing precision medicine.

## Introduction

Anxiety disorders (ANX) and major depression (MD) are the most prevalent psychiatric conditions, making a substantial contribution to poor health and disability globally (1). Both ANX and MD are more prevalent among women (1–3). Comorbidity between the two disorders occurs in over half of all cases (2, 3), and is associated with greater morbidity, worse treatment response, and higher suicide risk (4–6). Overlapping symptomatology, causal relationships, and shared environmental risk factors (7–9), as well as shared genetic risk and neurobiology, may explain the high comorbidity (10, 11).

Both ANX and MD exhibit moderate heritability (12, 13), and consistently show high genetic correlations ranging between 0.78 and 0.91 (14–16). However, comorbidity complicates the interpretation of these genetic correlations. Large genome-wide association studies (GWAS) for ANX do not exclude cases of MD, and *vice versa* (14–19). Polygenic risk scores (PRS) derived from these GWASs exhibit strong associations with comorbid ANX and MD cases (20). We previously reported that the PRS of MD was a stronger predictor of ANX than the PRS of ANX (10). While including comorbidity can enhance GWAS power for locus discovery (21), it may conflate genetic signals specific to ANX and MD, hampering the identification of distinct, disorder-specific, biological mechanisms (22). Stratifying ANX and MD by comorbidity status may elucidate their unique and shared biological mechanisms to inform tailored treatments. A critical step toward this goal is obtaining GWAS of ANX and MD that are stratified by comorbidity status (20, 21).

Here, we investigated the genetic relationship between ANX and MD using the UK Biobank for discovery GWAS (23), and the Norwegian Mother, Father and Child Study (MoBa) cohort for out-of-sample PRS prediction of diagnoses and symptoms (24). We removed comorbid ANX-MD cases, conducted ANX-only and MD-only GWAS of the UK Biobank sample, and compared them with GWAS of ANX and MD that included comorbid cases. We investigated the genetic relationship between ANX-only and MD-only compared to ANX-comorbid and MD-comorbid, and how their respective PRS associated with ANX and MD diagnoses and symptoms in MoBa. We also explored differences in biological pathways and how comorbidity affected causal relationships between ANX and MD using Mendelian Randomization (MR) (25, 26).

## Methods and Materials

We used two large, genotyped cohorts with data on relevant psychiatric phenotypes among Europeans for training and testing: the UK Biobank (UKB, N=500,000; Project No. 27412) for GWAS (Training; Supplement 1: Methods), and the Norwegian Mother, Father, and Child Cohort (MoBa, N=130,992) for independent testing (Testing; Supplement 1: Methods). These cohorts are linked to hospital diagnoses using ICD codes.

### Training dataset

We used UKB data to perform GWAS for anxiety disorders (ANX) without comorbid major depression (MD) (ANX-only; cases = 9,980 vs. controls = 179,442) and MD without comorbid ANX (MD-only; cases = 15,301 vs. controls = 179,038).

#### Diagnoses

The diagnoses, which are based on ICD-10 codes, were from hospital records of specialist healthcare services in the UK. Individuals with history of diagnoses of organic mental disorders (F00 – F09), psychotic disorders (F20 – F29), bipolar disorders (F30.0 – F30.9 and F31.0 – F31.9), or mixed anxiety and depressive disorder (F41.2) were excluded from analyses. ANX-only cases were diagnosed with agoraphobia (F40.0), social phobia (F40.1), panic disorder (F41.0), generalized anxiety disorder (F41.1), or other anxiety disorders (F41.3 - F41.9), excluding individuals with MD-only diagnoses. MD-only cases had a diagnosis of any depressive episode (F32.0 - F32.9), or recurrent depressive disorder (F33.0 - F33.9), excluding individuals with ‘ANX-only’ diagnoses. Controls for ANX-only (n=179,442) and MD-only (n=179,038) were randomly selected and did not overlap. Individuals with MD were excluded from the ANX-only GWAS control group, and individuals with ANX were excluded from the MD-only GWAS control group.

### Comparison GWAS data

We used publicly available GWAS data, excluding MoBa cohort data, for ANX with comorbid MD (ANX-comorbid; N_cases_=112,919, N_controls_=645,271) (15). Also, MD with comorbid ANX (MD-comorbid; N_cases_=403,025, N_controls_=1,538,479), excluding samples from the UKB and MoBa (19). These comorbidity-inclusive GWAS datasets were compared to the ANX-only and MD-only GWAS datasets (Supplement 1: Methods).

### Testing dataset

We obtained genotype and phenotype data on mothers and fathers (N=130,992) who participated in MoBa (24, 27). All study participants provided written informed consent upon recruitment, and individuals who withdrew during follow-up were excluded from our analysis. The Regional Committee for Medical and Health Research Ethics approved the present study (2016/1226/REK Sør-Øst C).

#### Diagnoses

Data on diagnoses of mental disorders were obtained from the Norwegian Patient Registry (2008–2022), which comprises specialist psychiatric healthcare diagnoses based on ICD-10 codes. We excluded individuals who withdrew their consent (n=484), with missing covariates (n=1,590), were related or had a diagnosis of organic mental disorders (F00 – F09), psychotic disorders (F20 – F29), or bipolar disorders (F30.0 – F30.9 and F31.0 – F31.9) (Supplement 1: Methods).

We grouped cases within MoBa cohort into three disorder-specific subgroups: 1. ANX-only (cases: 1,992, controls: 85,851) with the same criteria as in the UKB (excluding individuals with MD). 2. MD-only (cases: 7,486, controls: 83,146) using the same criteria as in the UKB (excluding individuals with ANX). 3. ANX-MD (cases:3,468, controls: 85,107) comprising individuals with both MD and ANX or mixed anxiety and depressive disorder (F41.2). The number of cases in each diagnostic category and controls, and reasons for exclusion, are provided in Supplement 1 (Figure S1).

#### Depression and anxiety symptoms

Symptoms of anxiety and depression were assessed using the validated 8-item Symptom Check List (SCL-8), which correlates highly with the original 25-item Hopkins Symptom Checklist (HSCL-25) (28). Depression items included feeling blue, feeling everything as an effort, worry, and hopelessness, while anxiety items comprised feeling scared, tense, nervous, and fearful. Responses were rated on a Likert scale, 1 – not bothered, 2 – a little bothered, 3 – quite bothered, and 4 - very bothered, reflecting how much respondents’ level of distress in the preceding two weeks. Pregnant women participating in MoBa (n = 67,219) completed the SCL-8 during the 30^th^ week of gestation, and this questionnaire was not administered to the fathers. After excluding individuals with missing covariates (n=787) and related individuals (n=11,570), the analysis included 54,862 mothers (Figure S2 in Supplement 1).

### Genetic analysis

#### Genotype data

*UKB*: DNA was extracted from blood samples and genome-wide genotyping was performed using the UK BiLEVE and UK Biobank Axiom arrays. Genotype imputation and centralized quality control procedures for the UKB genotypes are described in detail elsewhere (23).

*MoBa:* The Norwegian Institute of Public Health oversaw the blood sample storage and DNA extraction (https://www.fhi.no/en/publ/2012/protocols-for-moba/). Genotyping was performed using genome-wide arrays, and the genetic data were imputed and processed for quality control using a family-based pipeline (https://github.com/psychgen/MoBaPsychGen-QC-pipeline) (27). We limited our analyses to data from individuals of European ancestry.

#### Genome-wide association studies (GWAS)

We performed ANX-only and MD-only GWAS using version 3 of the UKB genetic data among individuals of “white British” ancestry classified according to self-declared ethnicity and genetic principal component (PC) analysis (as defined by UKB data field 22006). We conducted the analyses using REGENIE v3.4.1, applying an approximate Firth logistic regression (29). This approach allows the inclusion of related individuals in the analysis and efficiently handles samples with unbalanced case-control ratios. Age, sex, and the first 20 genotype PCs were included as covariates. Variants with an imputation INFO score < 0.8, minor allele count < 20, and non-autosomal variants were excluded from the association testing, resulting in 19,201,831 variants tested in ANX-only and 19,238,272 variants tested in MD-only GWAS.

#### Polygenic risk score (PRS)

PRSs were derived from ANX-only and MD-only GWAS. In addition, PRSs were derived from GWAS of ANX and MD, inclusive of comorbid cases, i.e., ANX-comorbid (15) and MD-comorbid (19). To obtain unbiased and maximally variable PRS estimates, we applied the PRS-PCA approach (30–32). Using PRSice (v2.3.3) (33), we calculated PRSs for a set of *p*-value cut-offs (1.0e-06, 1.0e-05, 1.0e-04, 1.0e-03, 1.0e-02, 5.0e-02, 1.0e-01, 5.0e-01, 1.0) (30). Then, we extracted the first principal component (PC1) from all PRSs across the *p*-value cut-offs. The PRSs were used to predict phenotypes in MoBa samples, applying PC1 in regression models. PRS were standardized, and effect estimates are reported per 1 SD increase in the PRS.

### Statistical analysis

#### PRS associations

Logistic regression was used to test the association between each of the four PRS, and each diagnostic category (i.e., ANX-only, MD-only, and ANX-MD) in case-control comparisons. Additionally, logistic regression was used in case–case comparisons to examine whether each PRS was differentially associated with one diagnostic category versus another.

Ordinal logistic regression was used to test associations between each PRS and depression and anxiety symptom levels. As a sensitivity analysis, linear regression models were also fitted. All models were adjusted for age, sex, and the first 10 genotype PCs. Using PLINK2 (version 2.00a3.7), we applied the relationship pruning algorithm (--king-cutoff 0.05) to exclude a member from related pairs of participants with a kinship coefficient greater than 0.05, preferentially retaining cases. We limited genetic relatedness analysis to SNPs that were directly genotyped in one or more genotyping batches with high imputation quality (INFO score > 0.97). Only one of the related individuals with a kinship coefficient greater than 0.05 was kept, prioritizing cases.

Effect sizes were represented as odds ratios and β values with 95% confidence intervals for the logistic and linear regressions, respectively. Benjamini-Hochberg correction was applied to *p*-values to account for 12 tests of the diagnoses and 32 tests of symptoms. We compared regression coefficients between models using a two-sample Z-test adjusted for shared controls. The test statistic was calculated as: 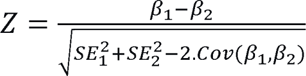, where the covariance term accounts for dependencies arising from the shared control group. Two-tailed *p*-values were derived from the standard normal distribution.

#### Heritability and Genetic Correlations

Linkage disequilibrium score regression (LDSC) (34) was applied to all four GWAS summary data to estimate genetic correlations between ANX and MD with or without comorbid cases. LDSC genetic-correlation analyses used LD scores computed from 489 unrelated European-ancestry individuals in the 1000 Genomes Project Phase 3, (https://doi.org/10.5281/zenodo.10515792). Single-nucleotide polymorphism (SNP) heritability for each phenotype was also computed. The heritability on the liability scale was estimated assuming the lifetime risk for ANX and MD to be 20% and 15%, respectively (15, 19).

#### Mendelian Randomization (MR)

We applied MR analyses using the R package TwoSampleMR (35) to examine causal associations between ANX and MD. These analyses were performed following the STROBE-MR guidelines (36). We applied inverse variance weighted (IVW) to highlight causal associations between ANX and MD (25). Additionally, weighted median (37), MR-Egger (38), and Causal Analysis Using Summary Effect estimates (CAUSE) (26) were applied as sensitivity analyses, and to account for confounding due to horizontal pleiotropy. Instruments were selected at a *p*-value threshold of 1e-05 to account for the low power of the ANX-only and MD-only GWASs (39), following previous studies’ application of a more relaxed p-value threshold for low-power GWASs (40, 41). The strength and heterogeneity of genetic instruments were assessed from F-statistics, Cochran’s Q, and I^2^ (42, 43). These analyses were performed using TwoSampleMR (version 0.5.8), and MRPRESSO (version 1.0). The resulting p-values were corrected using the Benjamini-Hochberg method for the 16 tests.

### Enrichment of Genes and Pathways

To identify and compare the biological mechanisms of ANX and MD, we employed GSA-MiXeR (v2.2.1), which can provide more specific genes and pathways relevant to a given phenotype (44). GSA-MiXeR estimates gene-level heritability for individual genes from the respective GWAS summary statistics. Fold enrichment for each gene can then be computed by calculating the ratio of the gene-level heritability estimated by the full model and the gene-level heritability estimated using a simplified baseline model, which assumes that genetic effects are evenly spread across the genome (44). GSA-MiXeR defines gene sets derived from GO terms of MsigDB v7.5 (biological processes, cellular component, and molecular function) and SynGO database (syngoportal.org). Enrichment of genes and pathways with Akaike Information Criterion (AIC) value greater than zero was considered reliable. We compared the enriched genes for ANX-only and MD-only as well as ANX-comorbid and MD-comorbid based on AIC and fold enrichment values.

## Results

### Sample characteristics

*UKB data:* ANX-only GWAS cases comprised 65% females and had a mean age (SD) of 58.3 (7.9) years, while controls, comprising of 53% females, had a mean age (SD) of 56.8 (8.0) years. Similarly, the MD-only GWAS cases comprised 62% females and had a mean age of 56.3 (7.9) years, whereas the controls were 53% females and the mean age (SD) was 56.8 (8.0) years. *MoBa data:* The proportions of females in MD-only, ANX-only, and ANX-MD cases was 70%, 81%, and 77%, respectively. The mean age (SD) for the corresponding case categories was 50.2 (5.7), 48.9 (5.3), and 49.2 (5.6) years, respectively. The control group comprised 57% females and had a mean age (SD) of 51.2 (5.4) years. For the prediction of symptoms of anxiety and depression, the sample included 54,862 mothers with a mean age (SD) of 30.1 (4.6) years.

### PRS associations with diagnoses in MoBa

To investigate the impact of removing comorbid cases from the GWAS in the training sample (UKB), we compared the associations of PRS from the different GWAS in the test sample (MoBa) (Table 1, Figure 1). MD-comorbid PRS and ANX-comorbid PRS showed the strongest associations with ANX-MD cases. In contrast, MD-only PRS and ANX-only PRS were associated with ANX-MD cases to a similar degree as with MD-only and ANX-only cases, respectively. MD-comorbid, ANX-comorbid, and MD-only PRS each showed differential associations with ANX-only compared to MD-only cases. ANX-only PRS did not show differential association with any of the three disorders, possibly due to limited statistical power from the smaller ANX-only GWAS sample size.

**Table 1:**
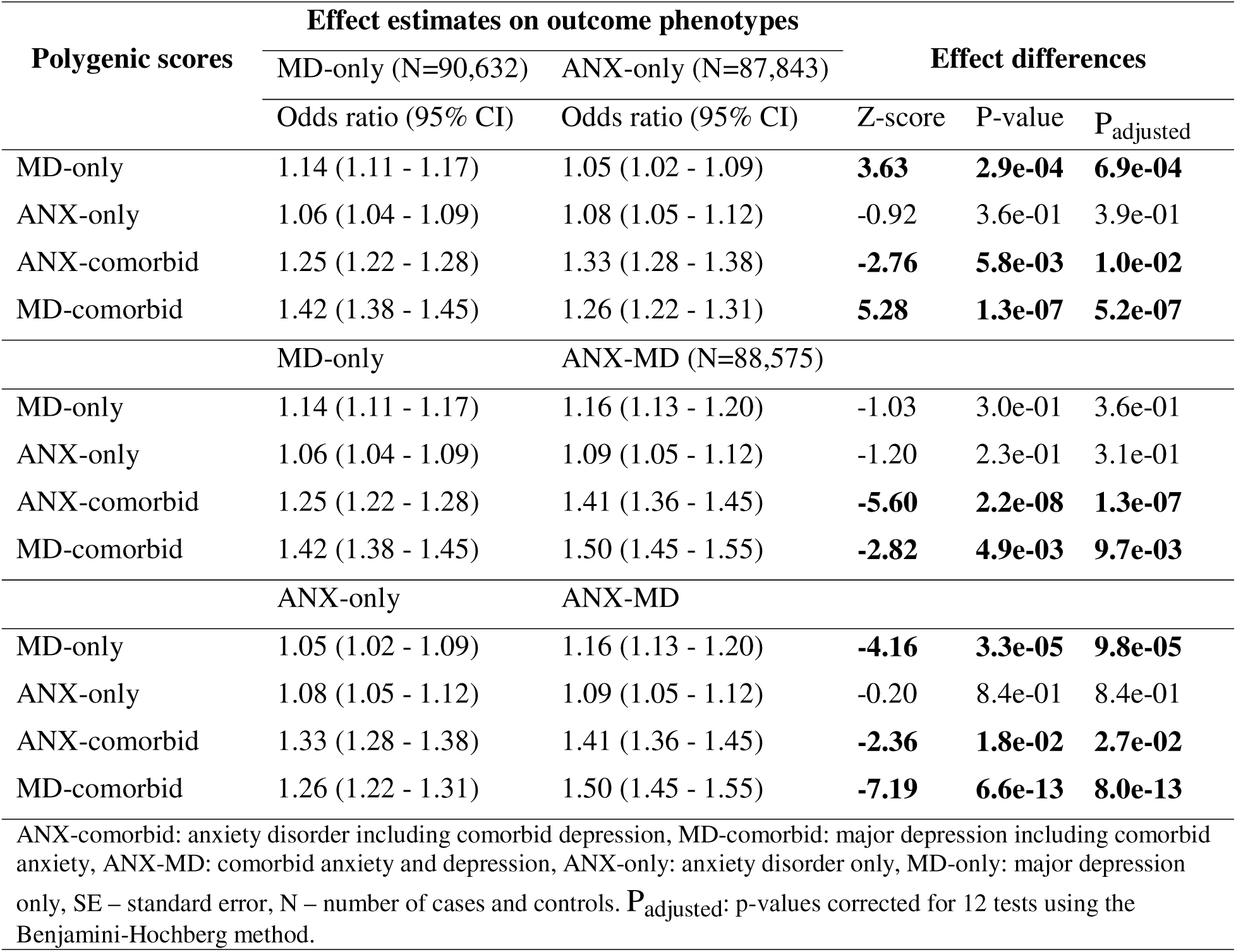
Comparison of the strength of associations of polygenic scores with phenotypes by comorbidity status in the MoBa cohort

**Figure 1.**
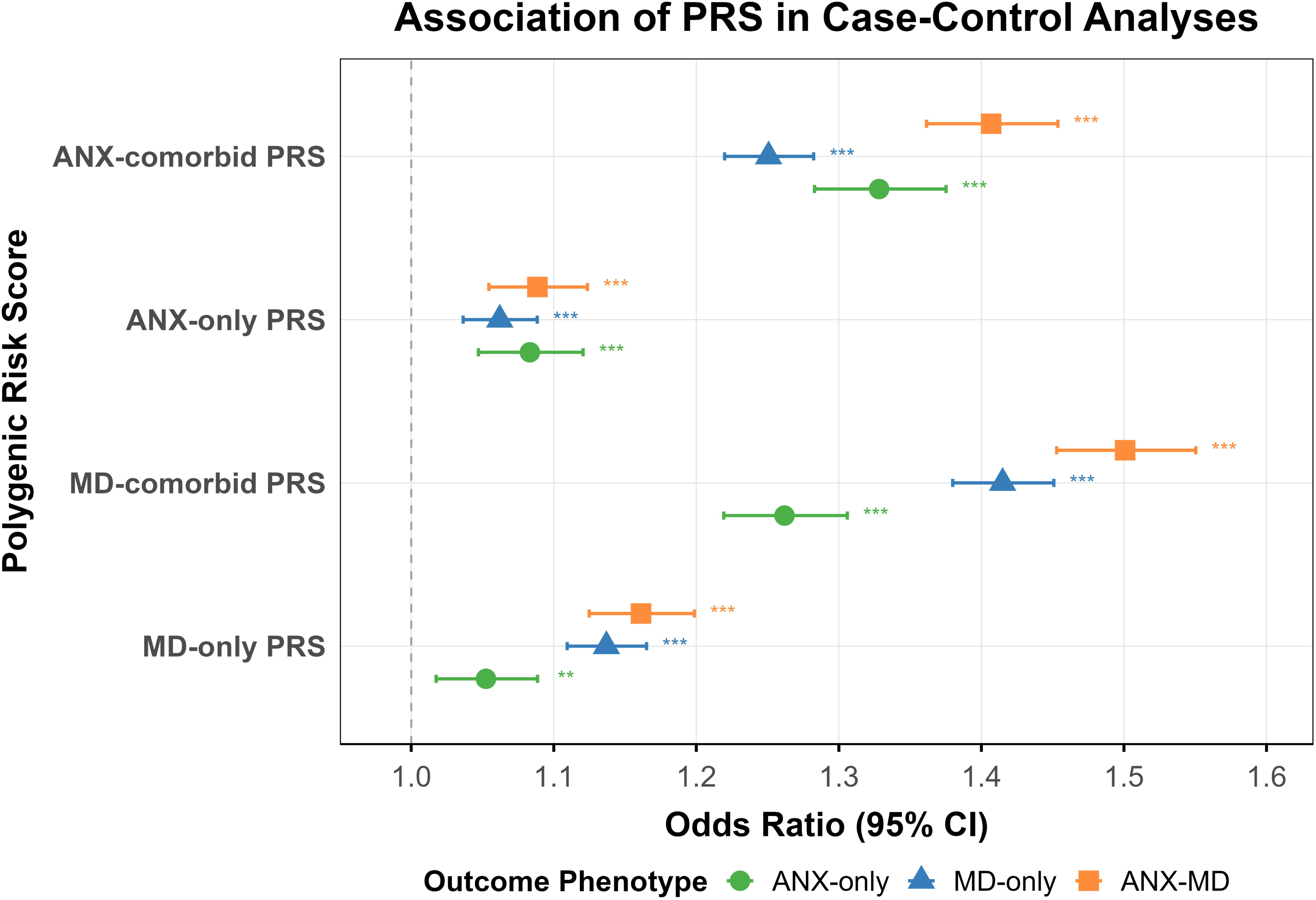
The association of PRS of differently defined anxiety and depressive disorders with anxiety disorder only (ANX-only), major depression only (MD-only), or comorbid anxiety and depression (ANX-MD) compared to controls. PRS: polygenic risk score; CI: confidence interval, ANX-comorbid: anxiety disorder including comorbid depression, MD-comorbid: major depression including comorbid anxiety. Odds ratios are displayed on the x-axis and PRS on the y-axis.

The patterns of PRS-diagnostic category associations depicted as differential associations in the case-case comparisons are consistent with those of the case-control analyses (Figure 2, and Table S1, Supplement 2).

**Figure 2.**
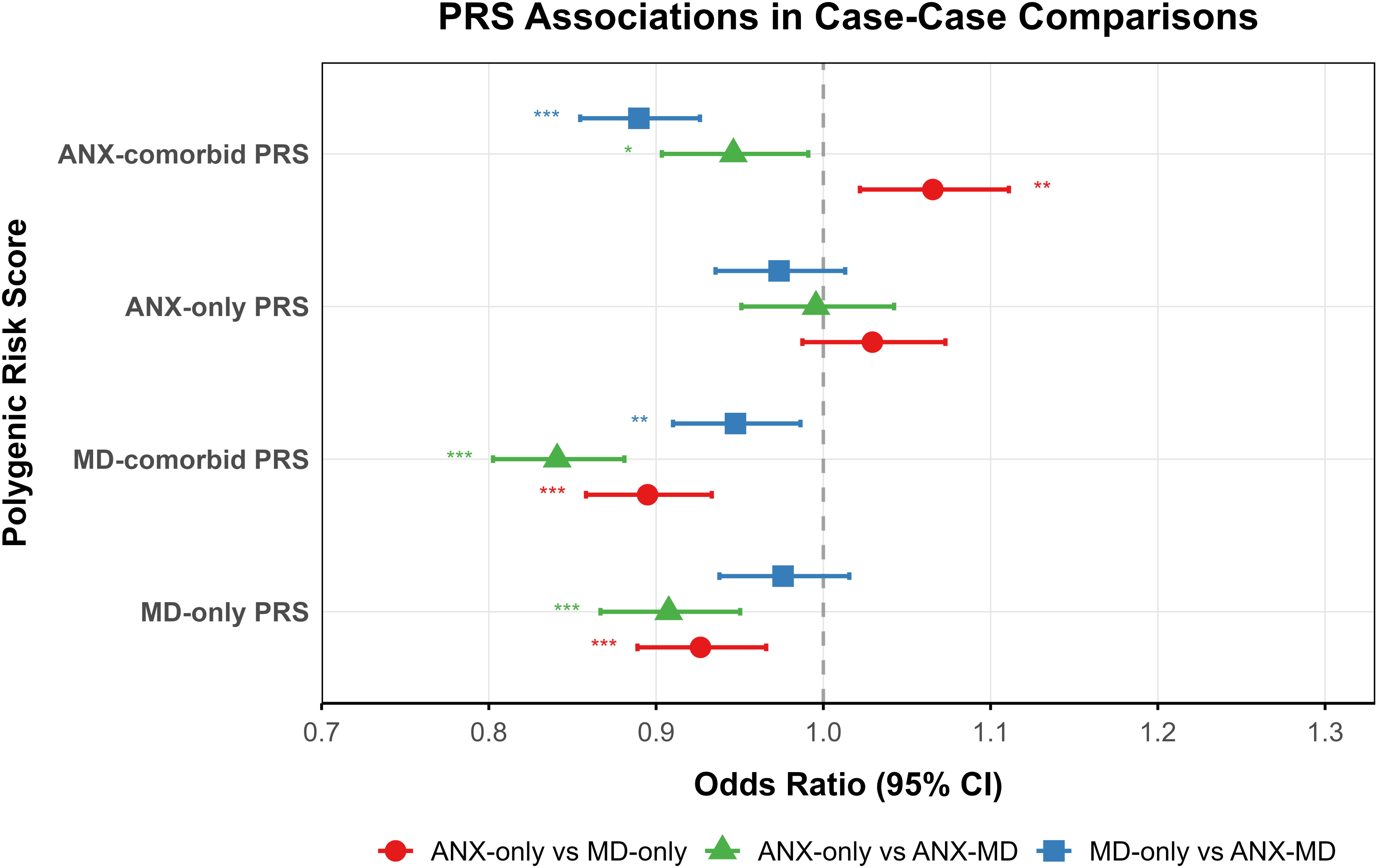
The association of PRS of differently defined anxiety and depressive disorders with anxiety disorder only (ANX-only), major depression only (MD-only), or comorbid anxiety and depression (ANX-MD) compared to each of the other diagnostic categories. PRS: polygenic risk score; CI: confidence interval, ANX-comorbid: anxiety disorder including comorbid depression, MD-comorbid: major depression including comorbid anxiety. Odds ratios are displayed on the x-axis and PRS on the y-axis.

### PRS associations with depression and anxiety symptoms

To investigate how comorbidity between ANX and MD affects genetic relationships beyond diagnostic categories, we performed ordinal logistic regression of symptom levels on the PRS from the four diagnostic categories. Each PRS showed significant associations with all eight symptoms of anxiety and depression after correction for multiple tests (p < 0.05), supporting the transdiagnostic nature of these symptoms (Figure 3). The strength of associations varied by PRSs and symptoms, particularly with MD-comorbid PRS generally showing larger effect sizes than ANX-only PRS (Table S2, Supplement 2), which is likely due to GWAS power. A sensitivity analysis applying linear regression models showed a similar pattern with all PRS exhibiting significant association with all symptoms (Table S3, Supplement 2).

**Figure 3.**
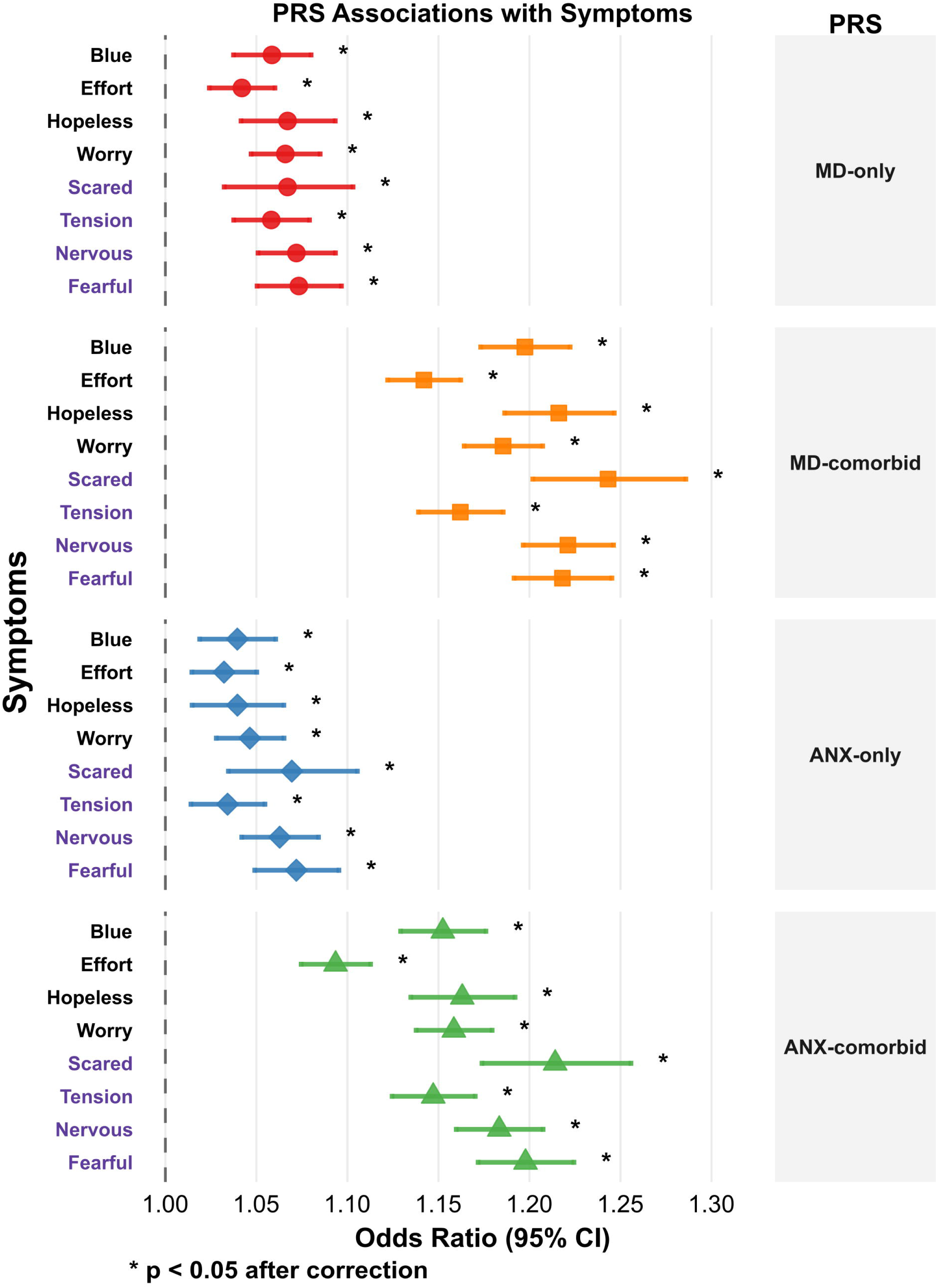
The association of polygenic scores (PRS) of differently defined anxiety and depressive disorders with symptoms of depression (Blue, Effort, Hopeless, Worry - black color) and symptoms of anxiety (Nervous, Fearful, Scared, Tension - purple color) reported by mothers from MoBa. ANX-comorbid: anxiety disorder including comorbid depression, MD-comorbid: major depression including comorbid anxiety, ANX-only: anxiety disorder only, and MD-only: major depression only.

**Figure 4.**
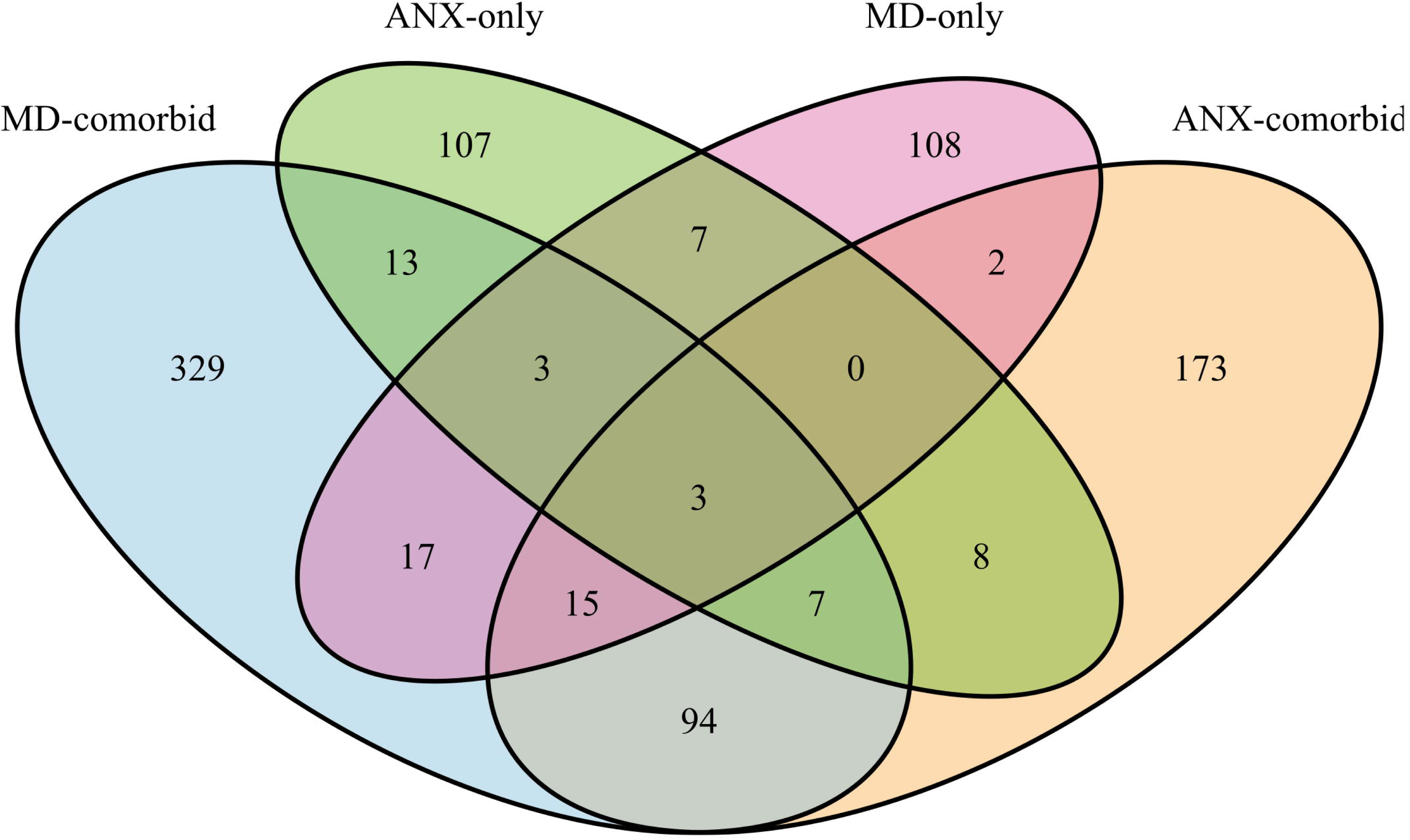
Venn diagram – The number of genes that showed enrichment in GSA-MiXeR analyses using GWAS data for different diagnoses. ANX-comorbid: anxiety disorder including comorbid depression, MD-comorbid: major depression including comorbid anxiety, ANX-only: anxiety disorder only, MD-only: major depression only.

### Heritability and Genetic Correlations

The observed (and liability) SNP heritability was 7.2% (13.1%) for MD-comorbid, 4.5% (11.6%) for ANX-comorbid, 10.5% (43.3%) for MD-only, and 5.4% (35.3%) for ANX-only. More conservative estimates of liability SNP heritability were 36.8% and 30.1%, respectively, assuming the lifetime risk for MD and ANX without comorbidity to be 60% of their overall prevalence. The genetic correlation between MD-comorbid and ANX-comorbid was higher (0.91; SE = 0.01, *p* < 5e-324) than between MD-only and ANX-only (0.53; SE = 0.11, *p* = 2.7e-06), illustrating that the inclusion of comorbid cases in the GWAS contributes to the high genetic correlation.

### Mendelian Randomization (MR)

In comorbidity-inclusive analyses, all methods supported bidirectional causal effects. For MD-comorbid on ANX-comorbid, IVW (β = 0.79; SE = 0.03, P_adjusted_ = 1.51e-157) was supported by the weighted median and MR-Egger. For ANX-comorbid on MD-comorbid, IVW (β = 0.55; SE = 0.03; P_adjusted_= 3.92e-75) supported by the weighted median, although MR-Egger showed attenuation (β = 0.09; SE = 0.08; P_adjusted_= 0.31), likely reflecting low power. CAUSE-MR confirmed bidirectional effects, with a larger effect of MD-comorbid on ANX-comorbid (γ = 0.54; 95% CI: 0.50-0.58; P_adjusted_= 1.02e-44) than the reverse (γ = 0.32; 95% CI: 0.30-0.35; P_adjusted_= 1.30e-29) (Table 2).

**Table 2.**
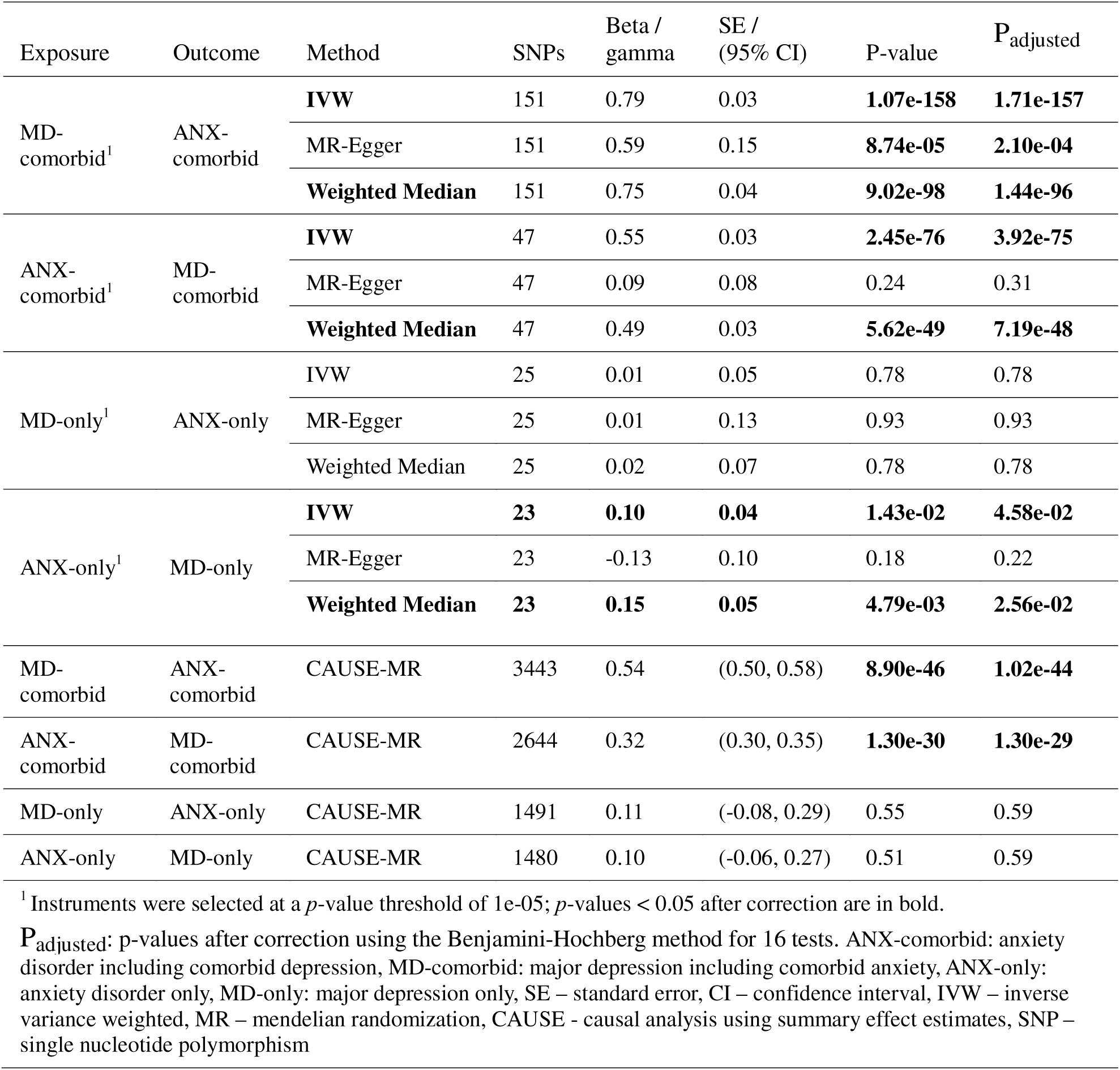
Bidirectional Mendelian Randomization Analyses of Anxiety Disorders and Major Depression

| Exposure | Outcome | Method | SNPs | Beta /<br>gamma | SE /<br>(95% CI) | P-value | P<sub>adjusted</sub> |
| --- | --- | --- | --- | --- | --- | --- | --- |
| MD-comorbid<sup>1</sup> | ANX-comorbid | **IVW** | 151 | 0.79 | 0.03 | **1.07e-158** | **1.71e-157** |
|  |  | MR-Egger | 151 | 0.59 | 0.15 | **8.74e-05** | **2.10e-04** |
|  |  | **Weighted Median** | 151 | 0.75 | 0.04 | **9.02e-98** | **1.44e-96** |
| ANX-comorbid<sup>1</sup> | MD-comorbid | **IVW** | 47 | 0.55 | 0.03 | **2.45e-76** | **3.92e-75** |
|  |  | MR-Egger | 47 | 0.09 | 0.08 | 0.24 | 0.31 |
|  |  | **Weighted Median** | 47 | 0.49 | 0.03 | **5.62e-49** | **7.19e-48** |
| MD-only<sup>1</sup> | ANX-only | IVW | 25 | 0.01 | 0.05 | 0.78 | 0.78 |
|  |  | MR-Egger | 25 | 0.01 | 0.13 | 0.93 | 0.93 |
|  |  | Weighted Median | 25 | 0.02 | 0.07 | 0.78 | 0.78 |
| ANX-only<sup>1</sup> | MD-only | **IVW** | **23** | **0.10** | **0.04** | **1.43e-02** | **4.58e-02** |
|  |  | MR-Egger | 23 | -0.13 | 0.10 | 0.18 | 0.22 |
|  |  | **Weighted Median** | **23** | **0.15** | **0.05** | **4.79e-03** | **2.56e-02** |
| MD-comorbid | ANX-comorbid | CAUSE-MR | 3443 | 0.54 | (0.50, 0.58) | **8.90e-46** | **1.02e-44** |
| ANX-comorbid | MD-comorbid | CAUSE-MR | 2644 | 0.32 | (0.30, 0.35) | **1.30e-30** | **1.30e-29** |
| MD-only | ANX-only | CAUSE-MR | 1491 | 0.11 | (-0.08, 0.29) | 0.55 | 0.59 |
| ANX-only | MD-only | CAUSE-MR | 1480 | 0.10 | (-0.06, 0.27) | 0.51 | 0.59 |

<sup>1</sup> Instruments were selected at a $p$ -value threshold of $1e-05$ ; $p$ -values $< 0.05$ after correction are in bold.
P<sub>adjusted</sub>: $p$ -values after correction using the Benjamini-Hochberg method for 16 tests. ANX-comorbid: anxiety disorder including comorbid depression, MD-comorbid: major depression including comorbid anxiety, ANX-only: anxiety disorder only, MD-only: major depression only, SE – standard error, CI – confidence interval, IVW – inverse variance weighted, MR – mendelian randomization, CAUSE - causal analysis using summary effect estimates, SNP – single nucleotide polymorphism

In comorbidity-exclusive analyses, IVW suggested a possible unidirectional effect of ANX-only on MD-only (β = 0.10; SE = 0.04; P_adjusted_= 0.045), supported by the weighted median (β = 0.15; SE = 0.05; P_adjusted_= 0.026), however, MR-Egger showed no evidence of an effect (P_adjusted_= 0.22). No significant effect was observed for MD-only on ANX-only with any method (Table 2). Instrument quality was lower for comorbidity-exclusive traits (F=19.5–24.4) than for comorbidity-inclusive traits (F=29–58; Table S4, Supplement 2). CAUSE-MR showed attenuated effects in both directions for ANX-only and MD-only (P_adjusted_ > 0.5). All analyses showed substantial heterogeneity (I² ≥ 0.95; Table S5, Supplement 2).

### Enrichment of Genes and Pathways

GSA-MiXeR identified significant fold enrichment for 148 genes in ANX-only and 155 in MD-only; Among these, 107 and 108 genes were unique to each condition, respectively, while 13 genes were enriched in both (*ERBB4, ACSL3, TPRG1, MAML3, KCNQ5, INSIG1, KCNB2, FER1L6, B3GLCT, IRX6, TSHZ3, TOX2*, and *FAM110A*). Enriched pathways for ANX-only included: microvesicle, CCR6 chemokine receptor binding, and cardiac muscle myoblast proliferation (Table S6, Supplement 2). For MD-only, pathways included negative regulation of myoblast proliferation, protein-O-linked fucosylation, and positive regulation of 3-UTR-mediated mRNA stabilization (Table S7, Supplement 2). A larger number of genes showed enrichment in ANX-comorbid (n=302) and MD-comorbid (n=481) than in ANX-only and MD-only, respectively. The proportion of overlapping genes (n=119) accounts for 39.5% of the maximum possible overlap (the gene set of 301 ANX-comorbid) (Tables S8 & S9, Supplement 2). In contrast, ANX-only and MD-only shared only 8.8% of the maximum possible overlap (the gene set of ANX-only). This difference in overlap proportions was significant (*p*-value < 0.0001) (Figure 3). Notably, the biological process interleukin 21 production was enriched in MD-only, ANX-comorbid, and MD-comorbid but not in ANX-only.

## Discussion

Here, we investigated the genetic relationship between ANX and MD using samples with and without comorbid cases. The genetic correlation between ANX and MD was significantly reduced after excluding comorbid cases. The ANX-comorbid PRS and MD-comorbid PRS showed the strongest associations with ANX-MD cases, suggesting that the genetic signal from the discovery GWAS is more representative of a comorbid phenotype than of ANX-only or MD-only. While MD-only PRS was differentially associated with MD-only cases compared with ANX-only cases, it also showed a significant association with ANX-MD and ANX-only relative to controls. Similarly, ANX-only PRS showed a significant association with ANX-only, MD-only, and ANX-MD cases, indicating pleiotropy. The MR results suggest a bidirectional causal association of ANX and MD, likely driven by comorbidity. These findings, together with the pattern of gene enrichment overlaps, suggest that MD-only and ANX-only may have more distinct underlying biological pathways than the shared biology inferred from comorbidity-inclusive GWAS.

ANX and MD without comorbidity showed only a moderate genetic correlation (r_g_ = 0.53) differs from the high genetic correlation observed for ANX and MD with comorbidity (r_g_ = 0.91). This latter finding aligns with previous GWAS that typically did not exclude comorbid cases (14–18). The heritability estimates for ANX-only and MD-only, which are comparable to or higher than those for their comorbidity-inclusive counterparts, align with evidence supporting higher heritability estimates for less heterogeneous phenotypes (45). These heritability estimates indicate that the ANX-only and MD-only GWAS were sufficiently powered for estimating genetic correlation. Our findings align with a recent family-based adoption study from Sweden, which showed a decrease in the genetic correlation from 0.83 to 0.46 after re-classification of cases into primary ANX or primary MD through diagnostic hierarchy (46). Consistent with its higher rates of comorbidity with MD relative to other anxiety phenotypes (47), generalized anxiety disorder also demonstrates elevated genetic correlations with MD (48).

The bidirectional causal associations between ANX-comorbid and MD-comorbid across multiple MR methods contrast with the absence of reliable causal effects between ANX-only and MD-only after rigorous sensitivity analyses. The ANX-comorbid and MD-comorbid GWAS were 10-fold and 22-fold larger than their comorbidity-exclusive counterparts, respectively, providing adequate power to detect causal associations even in the presence of significant instrument heterogeneity. The substantially smaller ANX-only and MD-only GWAS mean that even if true causal effects exist in the absence of comorbidity, our data may be inadequately powered to detect them. Overall, these findings underscore the importance of considering comorbidity when investigating shared genetic architecture and causal relationships between psychiatric disorders.

The trend for the strongest associations with comorbid cases (i.e., ANX-MD) for both MD-comorbid and ANX-comorbid PRS underscores the overrepresentation of comorbid cases in their respective GWAS. In line with our findings, a large study using biobank samples recently showed that the PRS of MD with comorbidity showed a stronger association with comorbid cases than with MD-only cases (20). Notably, MD-only and ANX-only PRS had similar associations with comorbid cases as with their respective disorders. Furthermore, ANX-comorbid, MD-comorbid, and MD-only PRS had significant differential associations with ANX-only cases compared to MD-only cases in the expected directions. Compared to healthy controls, all PRS were significantly associated with all three diagnostic categories, further evidence of genetic pleiotropy between ANX and MD. Similarly, the positive associations of all four PRS with all eight symptoms of ANX and MD, albeit to various degrees, underscore the transdiagnostic nature of these symptoms. This finding is consistent with the assertion that self-report symptoms indicate general dysphoria and could be associated with different disorder-specific PRS (49) as well as with significant phenotypic correlations across anxiety and depressive symptoms (50). Our findings may have potential clinical applications, e.g., sub-grouping patients with MD for treatment selection or predicting prognosis. Disorder-specific genetic risk may also have implications for predicting treatment outcomes (51, 52).

Gene enrichment analyses revealed that 39% of enriched genes overlapped between ANX and MD when comorbid cases were included, compared to only 9% when comorbid cases were excluded. It is important to conduct GWAS on well-defined MD phenotypes to identify MD-specific biological pathways (22). Consistent with this, interleukin-21 production is enriched in our gene sets for ANX-comorbid and MD with and without comorbidity, but not for ANX-only. These findings support the notion that ANX and MD have partially independent genetic underpinnings (20). While the greater proportion of overlapping genes may partly reflect differences in GWAS power, the inclusion of comorbid cases may also have contributed.

Efforts to improve genetic discoveries in complex phenotypes emphasize increasing sample sizes (53). Although a broad case definition can increase the power of GWAS (22), it can also increase phenotypic heterogeneity, hindering the discovery of subgroup-specific genetic loci essential for precision medicine (21). Thus, investigating the genetic relationships between disorders with pervasive comorbidity without accounting for comorbid cases may yield misleading results regarding both genetic architecture and functional annotations. The inclusion of comorbid cases in GWASs may have inflated the reported genetic overlap and shared loci between ANX and MD (10, 16). Current genome-wide approaches for ANX and MD might overlook some biological pathways related to only one of these disorders. As larger samples become available, GWAS for these sub-groups of patients is a logical next step to advancing precision medicine.

Our findings should be interpreted in the light of some limitations. Cases were defined using hospital diagnosis data and may include undetected comorbidity, and some controls may have undiagnosed ANX or MD. However, such misclassification would bias results toward the null, making our findings conservative. As different ANX subtypes have different comorbidity rates and genetic correlations with MD, the observed findings may partly reflect shifts in the proportion of specific ANX subtypes when comorbid cases are excluded. Differences in statistical power between training and testing samples may affect PRS effect estimates, particularly for ANX-only. Also, the ANX-only and MD-only GWAS were much smaller than those including comorbid individuals, which may have limited power to detect causal associations in MR. Similarly, variation in GWAS power may affect gene-set enrichment detection using GSA-MiXeR. Both the UKB and MoBa data used in these analyses are predominantly white European populations and of higher educational levels than the general populations, which need consideration in the interpretation of the findings.

In conclusion, our findings demonstrate that genetic liabilities for ANX and MD are more distinct than previously inferred from GWAS that included comorbid cases and support the existence of a comorbid group arising from true genetic pleiotropy. The results also implicate more specific underlying biological pathways across the ANX-MD spectrum. Larger GWAS of ANX-only and MD-only are needed to assess causal relationships and shared biological mechanisms in the absence of comorbidity. Together, genetic factors derived from ANX and MD, excluding comorbid cases, may facilitate precision medicine approaches and enable identification of biological mechanisms for tailored interventions.

## Data Availability

All data produced in the present study are available upon reasonable request to the authors.

## Acknowledgements

The authors received support from the Research Council of Norway (324252, 324499, 300309, 273291, 223273, 248778, 273446, 326813, 334920, 271555/F21, 324620), NordForsk (grant 156298), the European Economic Area and Norway Grants (EEA-RO-NO-2018-0535, EEA-RO-NO-2018-0573), the South-East Norway Regional Health Authority (2022-087 and 2022-073), The University of Oslo, KG Jebsen Stiftelsen (SKGJ-MED-021), the European Union’s Horizon 2020 research and innovation program (847776, 964874), and Marie Skłodowska-Curie grant (801133), Wellcome Leap, CARE Program (“FEMA-AD”), and the US National Institutes of Health (R01MH123724-01, R01MH124839-02, U24DA041123, R01AG076838, U24DA055330, and OT2 HL161847). This work was supported by the National Institutes of Health grants R01MH125938 (MT). The content is solely the responsibility of the authors and does not necessarily represent the official views of the funding agencies and the National Institutes of Health. The Norwegian Mother, Father and Child Cohort Study is supported by the Norwegian Ministry of Health and Care Services and the Ministry of Education and Research. This research is part of the HARVEST collaboration, supported by the Research Council of Norway (grant 229624). The Norwegian Centre for Mental Disorders Research (NORMENT) provided genotype data, funded by the Research Council of Norway (grant 223273), South East Norway Health Authorities, and Stiftelsen Kristian Gerhard Jebsen. The Center for Diabetes Research, the University of Bergen, provided genotype data funded by the ERC AdG project SELECTionPREDISPOSED, Stiftelsen Kristian Gerhard Jebsen, the Trond Mohn Foundation, the Research Council of Norway, the Novo Nordisk Foundation, the University of Bergen, and the Western Norway Health Authorities.

We are grateful to all individuals who participated in the Norwegian MoBa study. We thank the Norwegian Institute of Public Health for generating high-quality genomic data. We also thank the research participants and employees of the Psychiatric Genomic Consortium (Anxiety Disorder Working Group and Major Depressive Disorder Working Group) and UK Biobank. This work was performed on the TSD (Tjeneste for Sensitive Data) facilities, owned by the University of Oslo, operated and developed by the TSD service group at the University of Oslo, IT-Department (USIT). This research has been conducted using the UK Biobank Resource under Application Number 27412.

This work was previously made available as a preprint on the medical archive (medRxiv), https://doi.org/10.1101/2024.11.19.24317523

## Data availability

The comorbidity-inclusive GWAS summary statistics can be obtained from the Psychiatric Genomics Consortium (PGC) through requests made at https://pgc.unc.edu/for-researchers/data-access-committee/data-access-portal/. The comorbidity-exclusive GWAS summary statistics will become available on the GWAS Catalog - https://www.ebi.ac.uk/gwas/. The codes for GSA-MiXeR are publicly available at (https://github.com/precimed/gsa-mixer). The codes for Mendelian Randomization (https://mrcieu.github.io/TwoSampleMR/articles/introduction.html) and polygenic scores (https://github.com/deepchocolate/pgs/blob/main/pcPGS.R) are also in the public domain.

## References

1. Collaborators GBDMD (2022): Global, regional, and national burden of 12 mental disorders in 204 countries and territories, 1990-2019: a systematic analysis for the Global Burden of Disease Study 2019. Lancet Psychiatry. 9:137–150.

2. Penninx BW, Pine DS, Holmes EA, Reif A (2021): Anxiety disorders. Lancet. 397:914–927.

3. Kessler RC, Berglund P, Demler O, Jin R, Koretz D, Merikangas KR, et al. (2003): The epidemiology of major depressive disorder: results from the National Comorbidity Survey Replication (NCS-R). JAMA. 289:3095–3105.

4. Ter Meulen WG, Draisma S, van Hemert AM, Schoevers RA, Kupka RW, Beekman ATF, et al. (2021): Depressive and anxiety disorders in concert-A synthesis of findings on comorbidity in the NESDA study. J Affect Disord. 284:85–97.

5. Zhou Y, Cao Z, Yang M, Xi X, Guo Y, Fang M, et al. (2017): Comorbid generalized anxiety disorder and its association with quality of life in patients with major depressive disorder. Sci Rep. 7:40511.

6. Kalin NH (2020): The Critical Relationship Between Anxiety and Depression. Am J Psychiatry. 177:365–367.

7. Mathew AR, Pettit JW, Lewinsohn PM, Seeley JR, Roberts RE (2011): Co-morbidity between major depressive disorder and anxiety disorders: shared etiology or direct causation? Psychol Med. 41:2023–2034.

8. Thorp JG, Campos AI, Grotzinger AD, Gerring ZF, An J, Ong JS, et al. (2021): Symptom-level modelling unravels the shared genetic architecture of anxiety and depression. Nat Hum Behav. 5:1432–1442.

9. Wittchen HU, Beesdo K, Bittner A, Goodwin RD (2003): Depressive episodes--evidence for a causal role of primary anxiety disorders? Eur Psychiatry. 18:384–393.

10. Tesfaye M, Jaholkowski P, Shadrin AA, van der Meer D, Hindley GFL, Holen B, et al. (2024): Identification of novel genomic loci for anxiety symptoms and extensive genetic overlap with psychiatric disorders. Psychiatry Clin Neurosci. 78:783–791.

11. Hettema JM (2008): What is the genetic relationship between anxiety and depression? Am J Med Genet C Semin Med Genet. 148C:140–146.

12. Shimada-Sugimoto M, Otowa T, Hettema JM (2015): Genetics of anxiety disorders: Genetic epidemiological and molecular studies in humans. Psychiatry Clin Neurosci. 69:388–401.

13. Sullivan PF, Neale MC, Kendler KS (2000): Genetic epidemiology of major depression: review and meta-analysis. Am J Psychiatry. 157:1552–1562.

14. Purves KL, Coleman JRI, Meier SM, Rayner C, Davis KAS, Cheesman R, et al. (2020): A major role for common genetic variation in anxiety disorders. Mol Psychiatry. 25:3292–3303.

15. Strom NI, Verhulst B, Bacanu SA, Cheesman R, Purves KL, Gedik H, et al. (2026): Genome-wide association study of major anxiety disorders in 122,341 European-ancestry cases identifies 58 loci and highlights GABAergic signaling. Nat Genet. 58:275–288.

16. Als TD, Kurki MI, Grove J, Voloudakis G, Therrien K, Tasanko E, et al. (2023): Depression pathophysiology, risk prediction of recurrence and comorbid psychiatric disorders using genome-wide analyses. Nat Med. 29:1832–1844.

17. Howard DM, Adams MJ, Clarke TK, Hafferty JD, Gibson J, Shirali M, et al. (2019): Genome-wide meta-analysis of depression identifies 102 independent variants and highlights the importance of the prefrontal brain regions. Nat Neurosci. 22:343–352.

18. Levey DF, Gelernter J, Polimanti R, Zhou H, Cheng Z, Aslan M, et al. (2020): Reproducible Genetic Risk Loci for Anxiety: Results From approximately 200,000 Participants in the Million Veteran Program. Am J Psychiatry. 177:223–232.

19. Adams MJ, Streit F, Meng X, Awasthi S, Adey BN, Choi KW, et al. (2025): Trans-ancestry genome-wide study of depression identifies 697 associations implicating cell types and pharmacotherapies. Cell. 188:640–652 e649.

20. Coombes BJ, Landi I, Choi KW, Singh K, Fennessy B, Jenkins GD, et al. (2023): The genetic contribution to the comorbidity of depression and anxiety: a multi-site electronic health records study of almost 178 000 people. Psychol Med. 53:7368–7374.

21. Crouch DJM, Bodmer WF (2020): Polygenic inheritance, GWAS, polygenic risk scores, and the search for functional variants. Proc Natl Acad Sci U S A. 117:18924–18933.

22. Cai N, Revez JA, Adams MJ, Andlauer TFM, Breen G, Byrne EM, et al. (2020): Minimal phenotyping yields genome-wide association signals of low specificity for major depression. Nat Genet. 52:437–447.

23. Bycroft C, Freeman C, Petkova D, Band G, Elliott LT, Sharp K, et al. (2018): The UK Biobank resource with deep phenotyping and genomic data. Nature. 562:203–209.

24. Brandlistuen RE, Kristjansson D, Alsaker E, Valen R, Birkeland E, Royrvik EC, et al. (2025): Cohort Profile Update: The Norwegian Mother, Father and Child Cohort (MoBa). Int J Epidemiol. 54.

25. Burgess S, Butterworth A, Thompson SG (2013): Mendelian randomization analysis with multiple genetic variants using summarized data. Genet Epidemiol. 37:658–665.

26. Morrison J, Knoblauch N, Marcus JH, Stephens M, He X (2020): Mendelian randomization accounting for correlated and uncorrelated pleiotropic effects using genome-wide summary statistics. Nat Genet. 52:740–747.

27. Corfield EC, Frei O, Shadrin AA, Rahman Z, Lin A, Athanasiu L, et al. (2022): The Norwegian Mother, Father, and Child cohort study (MoBa) genotyping data resource: MoBaPsychGen pipeline v.1. bioRxiv.2022.2006.2023.496289.

28. Tamb K, Røysamb E (2014): Selection of questions to short-form versions of original psychometric instruments in MoBa. Norsk Epidemiologi. 24:195 – 201.

29. Mbatchou J, Barnard L, Backman J, Marcketta A, Kosmicki JA, Ziyatdinov A, et al. (2021): Computationally efficient whole-genome regression for quantitative and binary traits. Nat Genet. 53:1097–1103.

30. Coombes BJ, Ploner A, Bergen SE, Biernacka JM (2020): A principal component approach to improve association testing with polygenic risk scores. Genet Epidemiol. 44:676–686.

31. Lu ZA, Ploner A, Birgegard A, Eating Disorders Working Group of the Psychiatric Genomics C, Landen M, Bulik CM, et al. (2026): Leveraging transdiagnostic genetic liability to psychiatric disorders to dissect clinical outcomes of anorexia nervosa. Mol Psychiatry. 31:1475–1484.

32. Patel Y, Shin J, Sliz E, Tang A, Mishra A, Xia R, et al. (2024): Genetic risk factors underlying white matter hyperintensities and cortical atrophy. Nat Commun. 15:9517.

33. Choi SW, O’Reilly PF (2019): PRSice-2: Polygenic Risk Score software for biobank-scale data. Gigascience. 8.

34. Bulik-Sullivan BK, Loh PR, Finucane HK, Ripke S, Yang J, Schizophrenia Working Group of the Psychiatric Genomics C, et al. (2015): LD Score regression distinguishes confounding from polygenicity in genome-wide association studies. Nat Genet. 47:291–295.

35. Hemani G, Zheng J, Elsworth B, Wade KH, Haberland V, Baird D, et al. (2018): The MR-Base platform supports systematic causal inference across the human phenome. Elife. 7.

36. Skrivankova VW, Richmond RC, Woolf BAR, Yarmolinsky J, Davies NM, Swanson SA, et al. (2021): Strengthening the Reporting of Observational Studies in Epidemiology Using Mendelian Randomization: The STROBE-MR Statement. JAMA. 326:1614–1621.

37. Bowden J, Davey Smith G, Haycock PC, Burgess S (2016): Consistent Estimation in Mendelian Randomization with Some Invalid Instruments Using a Weighted Median Estimator. Genet Epidemiol. 40:304–314.

38. Bowden J, Davey Smith G, Burgess S (2015): Mendelian randomization with invalid instruments: effect estimation and bias detection through Egger regression. Int J Epidemiol. 44:512–525.

39. Cheng W, Parker N, Karadag N, Koch E, Hindley G, Icick R, et al. (2023): The relationship between cannabis use, schizophrenia, and bipolar disorder: a genetically informed study. Lancet Psychiatry. 10:441–451.

40. Li F, Tang M, Hao C, Yang M, Pan Y, Lei P (2024): Brain imaging traits and epilepsy: Unraveling causal links via mendelian randomization. Brain Behav. 14:e70051.

41. Sanna S, van Zuydam NR, Mahajan A, Kurilshikov A, Vich Vila A, Vosa U, et al. (2019): Causal relationships among the gut microbiome, short-chain fatty acids and metabolic diseases. Nat Genet. 51:600–605.

42. Bowden J, Del Greco MF, Minelli C, Davey Smith G, Sheehan NA, Thompson JR (2016): Assessing the suitability of summary data for two-sample Mendelian randomization analyses using MR-Egger regression: the role of the I2 statistic. Int J Epidemiol. 45:1961–1974.

43. Sanderson E, Spiller W, Bowden J (2021): Testing and correcting for weak and pleiotropic instruments in two-sample multivariable Mendelian randomization. Stat Med. 40:5434–5452.

44. Frei O, Hindley G, Shadrin AA, van der Meer D, Akdeniz BC, Hagen E, et al. (2024): Improved functional mapping of complex trait heritability with GSA-MiXeR implicates biologically specific gene sets. Nat Genet. 56:1310–1318.

45. Tropf FC, Lee SH, Verweij RM, Stulp G, van der Most PJ, de Vlaming R, et al. (2017): Hidden heritability due to heterogeneity across seven populations. Nat Hum Behav. 1:757–765.

46. Kendler KS, Abrahamsson L, Ohlsson H, Sundquist J, Sundquist K (2022): An Extended Swedish Adoption Study of Anxiety Disorder and Its Cross-Generational Familial Relationship With Major Depression. Am J Psychiatry. 179:640–649.

47. Saha S, Lim CCW, Cannon DL, Burton L, Bremner M, Cosgrove P, et al. (2021): Co-morbidity between mood and anxiety disorders: A systematic review and meta-analysis. Depress Anxiety. 38:286–306.

48. Morneau-Vaillancourt G, Coleman JRI, Purves KL, Cheesman R, Rayner C, Breen G, et al. (2020): The genetic and environmental hierarchical structure of anxiety and depression in the UK Biobank. Depress Anxiety. 37:512–520.

49. Huang L, Tang S, Rietkerk J, Appadurai V, Krebs MD, Schork AJ, et al. (2024): Polygenic Analyses Show Important Differences Between Major Depressive Disorder Symptoms Measured Using Various Instruments. Biol Psychiatry. 95:1110–1121.

50. Vaccarino AL, Evans KR, Sills TL, Kalali AH (2008): Symptoms of anxiety in depression: assessment of item performance of the Hamilton Anxiety Rating Scale in patients with depression. Depress Anxiety. 25:1006–1013.

51. Dold M, Bartova L, Souery D, Mendlewicz J, Serretti A, Porcelli S, et al. (2017): Clinical characteristics and treatment outcomes of patients with major depressive disorder and comorbid anxiety disorders - results from a European multicenter study. J Psychiatr Res. 91:1–13.

52. Hicks PB, Sevilimedu V, Johnson GR, Tal IR, Chen P, Davis LL, et al. (2023): Factors Affecting Antidepressant Response Trajectories: A Veterans Affairs Augmentation and Switching Treatments for Improving Depression Outcomes Trial Report. Psychiatr Res Clin Pract. 5:131–143.

53. Abdellaoui A, Yengo L, Verweij KJH, Visscher PM (2023): 15 years of GWAS discovery: Realizing the promise. Am J Hum Genet. 110:179–194.

