## Supplementary File for "Comorbidity alters the genetic relationship between anxiety disorders and major depression"

*** Corresponding authors**

### **Methods:**

### **1. UK Biobank**

The UK Biobank is an extensive biomedical database comprising 500,000 individuals aged 40-69 years recruited from 2006 to 2010 from various regions across the UK. The participants consented to provide data on lifestyle, physical measures, and biological samples. The database includes genome-wide data, clinical diagnoses based on the International Classification of Diseases (ICD), and questionnaires (1).

**Genetic data**

DNA was extracted from blood samples and genome-wide genotyping was performed using the UK BiLEVE and UK Biobank Axiom arrays. Genotype imputation and quality control procedures for the UKB genotypes are described in detail elsewhere (1).

### **2. The Norwegian Mother, Father and Child Cohort (MoBa)**

MoBa participants were recruited from all over Norway from 1999-2009. The women agreed to participate in 41% of the pregnancies. The cohort includes 94,834 mothers, 75,229 fathers, and 113 ,632 children (2). All participants had provided informed consent. The current study is based on version 12 of the quality-assured data files released for research in January 2019. The establishment of MoBa and initial data collection was based on a license from the Norwegian Data Protection Agency and approval from The Regional Committees for Medical and Health Research Ethics. MoBa cohort is currently regulated by the Norwegian Health Registry Act. The current study was approved by The Regional Committees for Medical and Health Research Ethics (2016/1226/REK sør-øst C).

**Genetic Data**

The Norwegian Institute of Public Health oversaw the blood sample storage and DNA extraction (<https://www.fhi.no/en/publ/2012/protocols-for-moba/>) (3). Genotyping was performed using the Illumina Global Screening Array, HumanCoreExome and OmniExpress arrays. The genetic data were imputed and processed for quality control using a family-aware pipeline (<https://github.com/psychgen/MoBaPsychGen-QC-pipeline>) (4). We restricted our analyses to data from individuals of European ancestry.

### **3. Genome-wide Association Study (GWAS) Summary Data.**

#### **Anxiety Disorders**

The summary data for anxiety disorders (ANX-comorbid) were obtained from the latest GWAS from the Psychiatric Genomics Consortium (PGC) Anxiety Disorders Working Group (5). The cases comprised individuals of European ancestry who had a history of any of generalized anxiety disorder, social phobia, panic disorder, agoraphobia, or specific phobias from 36 cohorts. Cases excluded individuals with severe mental health conditions, including schizophrenia, autism, and intellectual disability, but not comorbid mood or other anxiety-related disorders. The control group had no history of anxiety disorders, mood disorders, or severe mental health conditions (5). To avoid overlap with the study population in the testing sample, a leave-N-out meta-analysis excluding two Norwegian populations—MoBa and HUNT—was requested and obtained. The GWAS summary statistics used in the current study, therefore, comprised 112,919 cases and 645,271 controls, with an effective sample size of 384,407.

#### **Major Depression**

The European ancestry summary data for major depression (MD-comorbid) were obtained from a recent GWAS meta-analysis by the PGC major depressive disorder working group (6). A leave-N-out meta-analysis excluding 23andMe and the MoBa cohort produced GWAS summary statistics comprising 403,025 cases and 1,538,479 controls, with an effective sample size of 1,277,454. Another leave-N-out GWAS summary dataset, additionally excluding the UK Biobank data, was generated for analyses to avoid sample overlap with ANX-comorbid. The GWAS cases were defined using different codes, including ICD-8, ICD-9, ICD-10, DSM-IV, DSM-IV-TR, and DSM-5. Cohort case definitions had excluded individuals with bipolar disorder, non-affective psychosis/schizophrenia, major depression related to substance use disorder, but not individuals with comorbid anxiety disorders. Exclusion criteria for the controls varied across cohorts; however, most excluded individuals had a history of major depression, bipolar disorder, or substance use disorder, as well as individuals who had scored high on symptom screening measures such as the CES-D.

### **Figure S1.** Flow chart showing the parents from MoBa for the prediction of anxiety and depressive disorders and the reasons for exclusion.

MoBa parents (n = 130,992)

Males: 53,358

Females: 77,634

Withdrew consent (n = 484)

Parents with consent

(n = 130,508)

Missing age data (n = 1,590)

Parents with covariate data

(n = 128,918)

Anxiety-only (ANX-only)

(Cases: 2,118; controls: 114,494)

Unrelated individuals

(Cases: 2,100; controls: 86,452)

Final sample

(Cases: 1,992; controls: 85,851)

n = 28,060*

n = 709**

Depression-only (MD-only)

(Cases:8,380; controls: 114,494)

Unrelated individuals

(Cases: 8,160; controls: 83,726)

Final sample

(Cases: 7,486; controls: 83,146)

n = 30,988*

n = 1,254**

Comorbid (ANX-MD)

(Cases: 3,926; controls: 114,494)

Unrelated individuals

(Cases: 3,869; controls: 85,703)

Final sample

(Cases: 3,468; controls: 85,107

n = 28,848*

n = 997**

* Excluded randomly one of a pair with kinship coefficient > 0.05; ** Excluded because of comorbid psychiatric disorder

### **Figure S2.** Flow chart showing study participant mothers analyzed for the prediction of symptoms of anxiety and depression and the reasons for exclusion. (N.B. Fathers were not included in this survey)

Participants with genetic data (n = 77,634)

Withdrew consent (n = 272)

Participants with consent (n = 77,362)

Missing data on symptoms of anxiety and depression (n = 10,143)

Participants with data on symptoms of anxiety and depression (n = 67,219)

Participants with age data (n = 66,432)

Final number of participants for analyses

(n = 54,862)

Missing age data (n = 787)

Randomly excluded one of a pair with kinship coefficient > 0.05 (n = 11,570)

### **References**

1.	Bycroft C, Freeman C, Petkova D, Band G, Elliott LT, Sharp K, et al. (2018): The UK Biobank resource with deep phenotyping and genomic data. *Nature*. 562:203–209.

2.	Brandlistuen RE, Kristjansson D, Alsaker E, Valen R, Birkeland E, Royrvik EC, et al. (2025): Cohort Profile Update: The Norwegian Mother, Father and Child Cohort (MoBa). *Int J Epidemiol*. 54.

3.	Paltiel L, Haugen A, Skjerden T, Harbak K, Bækken S, Strensrud NK, et al. (2014): The biobank of the Norwegian Mother and Child Cohort Study – present status. *Nor J Epidemiol*. 24:29–35.

4.	Corfield EC, Frei O, Shadrin AA, Rahman Z, Lin A, Athanasiu L, et al. (2022): The Norwegian Mother, Father, and Child cohort study (MoBa) genotyping data resource: MoBaPsychGen pipeline v.1. *bioRxiv*.2022.2006.2023.496289.

5.	Strom NI, Verhulst B, Bacanu SA, Cheesman R, Purves KL, Gedik H, et al. (2026): Genome-wide association study of major anxiety disorders in 122,341 European-ancestry cases identifies 58 loci and highlights GABAergic signaling. *Nat Genet*. 58:275–288.

6.	Adams MJ, Streit F, Meng X, Awasthi S, Adey BN, Choi KW, et al. (2025): Trans-ancestry genome-wide study of depression identifies 697 associations implicating cell types and pharmacotherapies. *Cell*. 188:640–652 e649.
